## Supplementary tables for "The association of neutralizing antibodies with protection against symptomatic dengue virus infection varies by serotype, prior infection history, and assay condition"

**Table S1. ROC analysis and logistic regression of NT _50_ magnitude as a predictor for developing a subsequent symptomatic disease, stratified by infection history and incoming serotype**.

|  | **Combined** | | | | | | |
| --- | --- | --- | --- | --- | --- | --- | --- |
|  | **AUC**^a^ | **CI 95**^b^ | **Threshold (NT_50_)** | **Sensitivity** | **Specificity** | **OR**^c^ | **CI 95** |
| **DENV1** |  |  |  |  |  |  |  |
| Part.Mature^d^ Vero | 0.74 | 0.50 to 0.99 | 325.90 | 1 | 0.44 | *ns* | |
| Mature Vero | 0.77 | 0.51 to 1 | 206.84 | 1 | 0.67 | *ns* | |
| Part.Mature^d^ Vero DC-SIGN | 0.82 | 0.61 to 1 | 227.00 | 1 | 0.44 | *ns* | |
| Mature Vero DC-SIGN | 0.68 | 0.42 to 0.94 | 148.35 | 0.9 | 0.44 | *ns* | |
| **DENV2** |  |  |  |  |  |  |  |
| Part.Mature^d^ Vero | 0.62 | 0.47 to 0.77 | 968.34 | 1 | 0.18 | 0.86 | 0.73 to 0.99 |
| Mature Vero | 0.71 | 0.57 to 0.85 | 441.75 | 0.91 | 0.5 | 0.69 | 0.52 to 0.85 |
| Part.Mature^d^ Vero DC-SIGN | 0.65 | 0.50 to 0.79 | 714.55 | 0.94 | 0.21 | 0.83 | 0.68 to 0.98 |
| Mature Vero DC-SIGN | 0.68 | 0.54 to 0.82 | 229.85 | 0.91 | 0.32 | 0.64 | 0.41 to 0.9 |
| **DENV3** |  |  |  |  |  |  |  |
| Part.Mature^d^ Vero | 0.52 | 0.52 to 0.85 | 281.35 | 0.85 | 0.46 | *ns* | |
| Mature Vero | 0.51 | 0.33 to 0.70 | 201.10 | 0.7 | 0.42 | *ns* | |
| Part.Mature^d^ Vero DC-SIGN | 0.61 | 0.44 to 0.78 | 304.75 | 0.85 | 0.31 | *ns* | |
| Mature Vero DC-SIGN | 0.64 | 0.47 to 0.81 | 148.10 | 0.9 | 0.27 | *ns* | |
|  | **Primary** | | | | | | |
|  | **AUC**^a^ | **CI 95**^b^ | **Threshold (NT_50_)** | **Sensitivity** | **Specificity** | **OR**^c^ | **CI 95** |
| **DENV1** |  |  |  |  |  |  |  |
| Part.Mature^d^ Vero | 0.74 | 0.50 to 0.99 | 325.90 | 1 | 0.44 | *ns* | *-* |
| Mature Vero | 0.77 | 0.51 to 1 | 206.84 | 1 | 0.67 | *ns* | *-* |
| Part.Mature^d^ Vero DC-SIGN | 0.82 | 0.61 to 1 | 227.00 | 1 | 0.44 | *ns* | *-* |
| Mature Vero DC-SIGN | 0.68 | 0.42 to 0.94 | 148.35 | 0.9 | 0.44 | *ns* | *-* |
| **DENV2** |  |  |  |  |  |  |  |
| Part.Mature^d^ Vero | 0.65 | 0.42 to 0.87 | 558.15 | 1 | 0.38 | *ns* | - |
| Mature Vero | 0.72 | 0.51 to 0.93 | 465.65 | 1 | 0.46 | 0.62 | 0.38 to 0.88 |
| Part.Mature^d^ Vero DC-SIGN | 0.70 | 0.50 to 0.90 | 351.50 | 0.94 | 0.38 | *ns* | - |
| Mature Vero DC-SIGN | 0.75 | 0.56 to 0.95 | 74.88 | 0.94 | 0.54 | 0.09 | 0.01 to 0.52 |
| **DENV3** |  |  |  |  |  |  |  |
| Part.Mature^d^ Vero | 0.73 | 0.52 to 0.93 | 273.55 | 0.93 | 0.33 | *ns* | *-* |
| Mature Vero | 0.49 | 0.26 to 0.72 | 240.10 | 0.79 | 0.17 | *ns* | *-* |
| Part.Mature^d^ Vero DC-SIGN | 0.59 | 0.37 to 0.82 | 532.60 | 0.93 | 0.17 | *ns* | *-* |
| Mature Vero DC-SIGN | 0.73 | 0.52 to 0.93 | 110.55 | 0.93 | 0.17 | *ns* | *-* |
|  | **Secondary** | | | | | | |
|  | **AUC**^a^ | **CI 95**^b^ | **Threshold (NT_50_)** | **Sensitivity** | **Specificity** | **OR**^c^ | **CI 95** |
| **DENV1** |  |  |  |  |  |  |  |
| Part.Mature^d^ Vero | - | - | - | - | - | - | - |
| Mature Vero | - | - | - | - | - | - | - |
| Part.Mature^d^ Vero DC-SIGN | - | - | - | - | - | - | - |
| Mature Vero DC-SIGN | - | - | - | - | - | - | - |
| **DENV2** |  |  |  |  |  |  |  |
| Part.Mature^d^ Vero | 0.60 | 0.38 to 0.81 | 981.84 | 1 | 0.2 | *ns* |  |
| Mature Vero | 0.68 | 0.48 to 0.89 | 475.83 | 1 | 0.47 | 0.72 | 0.5 to 0.95 |
| Part.Mature^d^ Vero DC-SIGN | 0.61 | 0.4 to 0.83 | 853.75 | 0.94 | 0.27 | *ns* |  |
| Mature Vero DC-SIGN | 0.66 | 0.45 to 0.87 | 455.50 | 0.94 | 0.33 | *ns* |  |
| **DENV3** |  |  |  |  |  |  |  |
| Part.Mature^d^ Vero | 0.51 | 0.16 to 0.86 | 810.73 | 0.67 | 0.14 | *ns* | *-* |
| Mature Vero | 0.59 | 0.23 to 0.97 | 23.18 | 1 | 0.07 | *ns* | *-* |
| Part.Mature^d^ Vero DC-SIGN | 0.55 | 0.18 to 0.91 | 693.55 | 0.67 | 0.07 | *ns* | *-* |
| Mature Vero DC-SIGN | 0.44 | 0.15 to 0.74 | 483.97 | 0.83 | 0.07 | *ns* | *-* |

^a^AUC = Area under the curve, ^b^CI95 = 95% Confidence interval, ^c^OR = Odds ratio, ^d^Part. Mature = Partially mature virion.
