## Supplementary Figures for "The association of neutralizing antibodies with protection against symptomatic dengue virus infection varies by serotype, prior infection history, and assay condition"

### Figure S1

A.

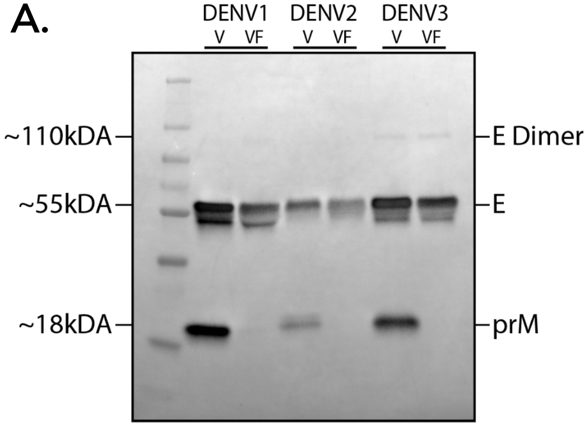


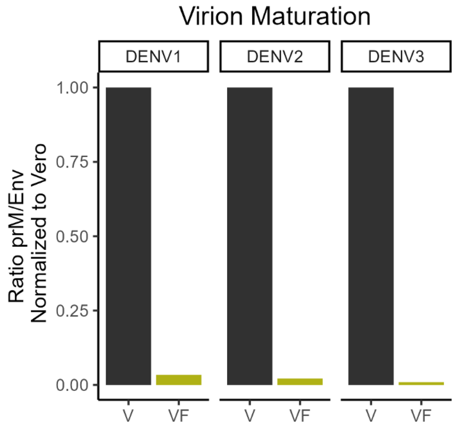


B.

| | | Copy genome (vRNA/mL) | Vero Viral Titer (FFU/mL) | Vero Infectivity (vRNA:FFU) | Vero DC-SIGN Viral Titer (FFU/mL) | Vero DC-SIGN Infectivity (vRNA:FFU) |
|---|---|---|---|---|---|---|
| DENV1 | Partially Mature | 5.55E+09 | 2.70E+06 | 2057 | 9.60E+06 | 578 |
| | Mature | 2.45E+09 | 4.30E+06 | 569 | 5.90E+06 | 415 |
| DENV2 | Partially Mature | 2.42E+10 | 1.13E+06 | 21386 | 1.90E+06 | 12719 |
| | Mature | 1.02E+10 | 1.58E+06 | 6428 | 3.30E+06 | 3078 |
| DENV3 | Partially Mature | 9.33E+09 | 7.87E+05 | 11859 | 2.73E+06 | 3417 |
| | Mature | 7.14E+09 | 1.92E+06 | 3729 | 6.43E+06 | 1112 |

C.

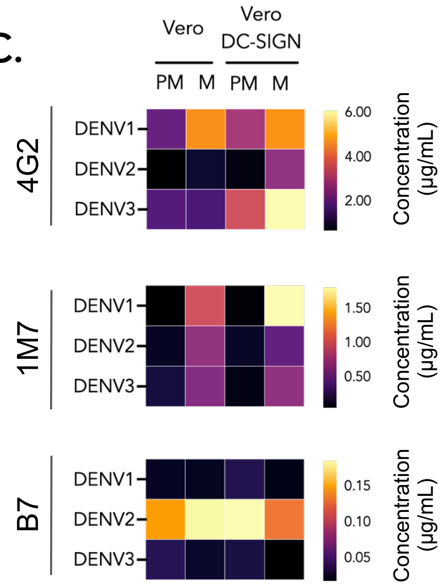


D.

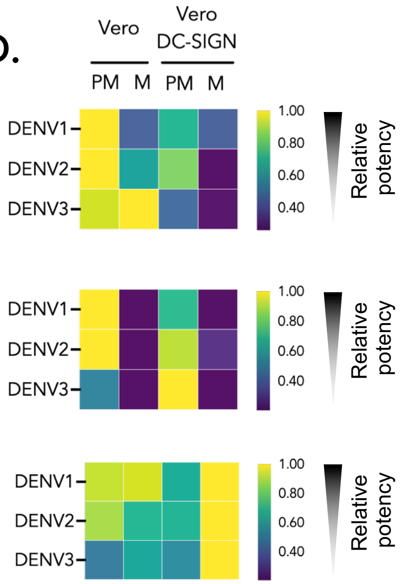

**Figure 1. Characterization of maturation state, infectivity, and mAb neutralization of viral stocks.** (A) Representative Western blot image (left) of DENV1-3 viral supernatants from Vero (V) and Vero-furin cells (VF) stained with anti-E and anti-prM antibodies. (B) Quantification of the increase in maturation state (prM/E) of DENV1-3 virions produced in VF cells compared to Vero cells (lower value = more mature) as seen by relative western blot band intensity. (C) Neutralization potency of monoclonal antibodies (mAbs), with $EC_{50}$ values in ug/mL displayed across assay conditions. The darker color corresponds to mAbs with the highest neutralization potency. (D) Relative neutralization potency of mAbs. Values are normalized by the lowest $EC_{50}$ observed among the four assay conditions (most potent), calculated as (1 / ($EC_{50}$ of assay condition X / lowest $EC_{50}$ of four assay condition)). Large color variation indicates a strong impact of change in assay conditions. Relative $EC_{50}$ value of 1 (yellow) indicates highest potency, of 0.2 (dark purple) least potent.

### Figure S2

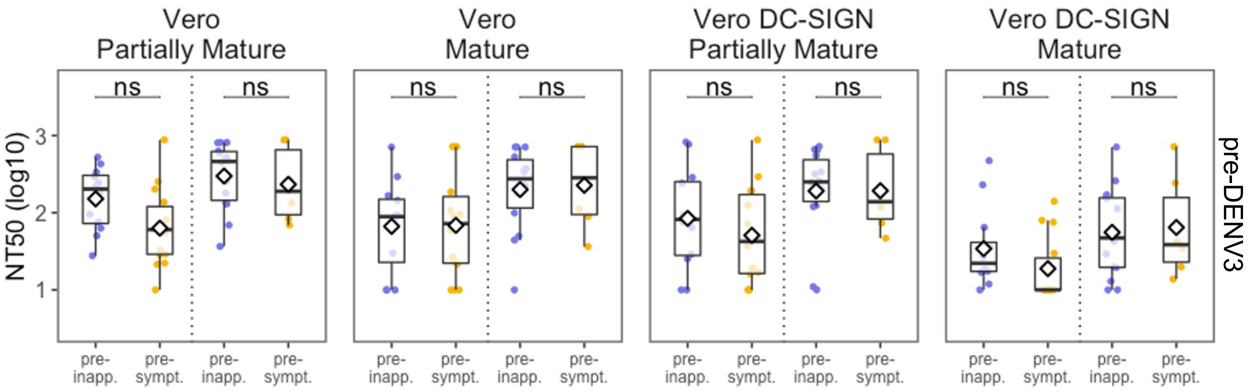


**Figure S2. Cross-reactive nAb titers of pre-inapparent and pre-symptomatic DENV3 infection participants stratified by infection history.** Asterisks indicate Benjamini Hochberg adjusted p-values for the Wilcoxon test. p-values: ns, >0.05; *, <0.05; **, <0.01; ***, <0.001; ****, <0.0001.

#### Figure S3

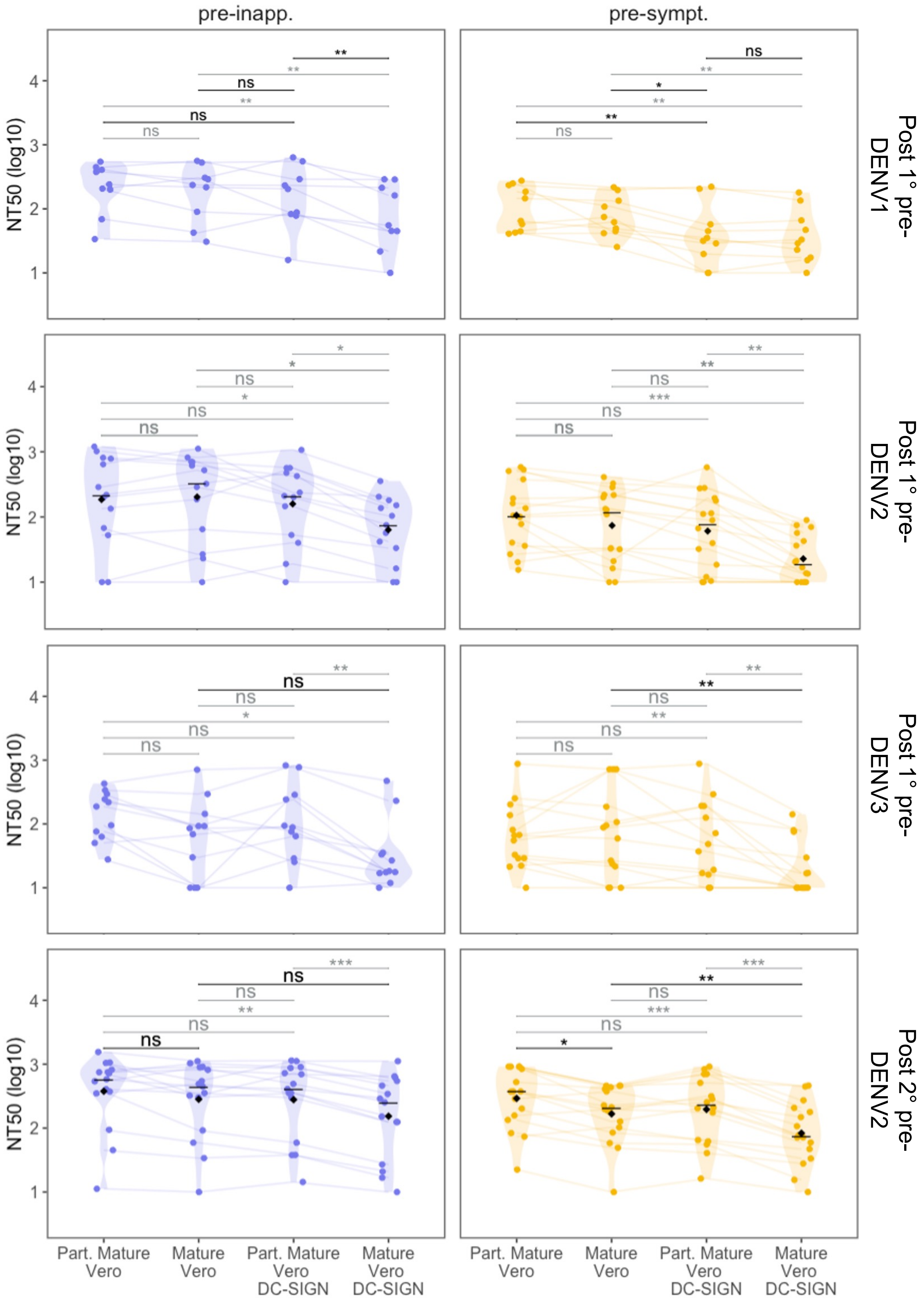

pre-inapp.
pre-sympt.
NT50 (log10)
Post 1° pre-DENV1
Post 1° pre-DENV2
Post 1° pre-DENV3
Post 2° pre-DENV2
Part. Mature Vero
Mature Vero
Part. Mature Vero DC-SIGN
Mature Vero DC-SIGN

**Figure S3. Impact of change in assay condition on the nAb response of pre-inapparent and pre-symptomatic infection participants stratified by infection history.** Post-1°: post-primary infection, Post-2°: post-secondary infection. Asterisks indicate Benjamini Hochberg adjusted p.values for paired Wilcoxon tests. p-values: ns, >0.05; *, <0.05; **, <0.01; ***, <0.001; ****, <0.0001.
